# The clinical characteristics of COVID-19: a retrospective analysis of 104 patients from the outbreak on board the Diamond Princess cruise ship in Japan

**DOI:** 10.1101/2020.03.18.20038125

**Authors:** Sakiko Tabata, Kazuo Imai, Shuichi Kawano, Mayu Ikeda, Tatsuya Kodama, Kazuyasu Miyoshi, Hirofumi Obinata, Satoshi Mimura, Tsutomu Kodera, Manabu Kitagaki, Michiya Sato, Satoshi Suzuki, Toshimitsu Ito, Yasuhide Uwabe, Kaku Tamura

## Abstract

**Background:** The ongoing outbreak of the coronavirus disease 2019 (COVID-19) is a global threat. Identification of markers for symptom onset and disease progression is a pressing issue. We compared the clinical features on admission among patients who were diagnosed with asymptomatic, mild, and severe COVID-19 at the end of observation.

**Methods:** This retrospective, single-center study included 104 patients with laboratory-confirmed COVID-19 from the mass infection on the Diamond Princess cruise ship from February 11 to February 25, 2020. Clinical records, laboratory data, and radiological findings were analyzed. Clinical outcomes were followed up until February 26, 2020. Clinical features on admission were compared among those with different disease severity at the end of observation. Univariate analysis identified factors associated with symptom onset and disease progression.

**Findings:** The median age was 68 years, and 54 patients were male. Briefly, 43, 41, and 20 patients on admission and 33, 43, and 28 patients at the end of observation had asymptomatic, mild, and severe COVID-19, respectively. Serum lactate hydrogenase levels were significantly higher in 10 patients who were asymptomatic on admission but developed symptomatic COVID-19 compared with 33 patients who remained asymptomatic throughout the observation period. Older age, consolidation on chest computed tomography, and lymphopenia on admission were more frequent in patients with severe COVID-19 than those with mild COVID-19 at the end of observation.

**Interpretation:** Lactate dehydrogenase level is a potential predictor of symptom onset in COVID-19. Older age, consolidation on chest CT images, and lymphopenia might be risk factors for disease progression of COVID-19 and contribute to the clinical management.

**Funding:** Not applicable.

**Research in context:** *Evidence before this study:* We searched the PubMed database from its inception until March 1, 2020, for articles published in English using the keywords “novel coronavirus,” “2019 novel coronavirus,” “2019-nCoV,” “Severe acute respiratory syndrome coronavirus 2,” “SARS-CoV2,” “COVID-19,” “mass infection,” “herd infection,” “cruise ship,” “Diamond Princess,” “asymptomatic,” and “subclinical.” There were no published clinical studies featuring COVID-19 as a result of mass infection on board a cruise ship. We found published articles entitled “Characteristics of COVID-19 infection in Beijing” and “Radiological findings from 81 patients with COVID-19 pneumonia in Wuhan, China: a descriptive study,” which compared patients with asymptomatic, mild, and severe COVID-19. However, none of the studies described potential markers for symptom onset or disease progression in patients with COVID-19.

*Added value of this study:* We present the differences in clinical characteristics of 104 patients with laboratory-confirmed COVID-19 as a result of mass infection on the Diamond Princess cruise ship who were treated at Self-Defense Forces Central Hospital, Japan from February 11 to February 25, 2020. On admission, 43, 41, and 20 patients had asymptomatic, mild, and severe COVID-19, respectively, whereas 33, 43, and 28 patients were determined to have asymptomatic, mild, and severe COVID-19, respectively, at the end of observation. During the observation period, 10 of the 43 (23.3%) asymptomatic patients on admission developed symptoms of COVID-19. Conversely, eight of the 84 (9.5%) patients with asymptomatic and mild COVID-19 on admission developed severe disease during the observation period. The serum lactate dehydrogenase (LDH) levels were significantly higher in 10 patients who were initially asymptomatic on admission to the hospital and developed symptomatic COVID-19 during the observation period compared with 33 patients who remained asymptomatic throughout the observation period. The prevalence rates of consolidation on chest computed tomography (CT) images and lymphopenia were significantly higher in eight patients who developed severe COVID-19 during the observation period compared with the 76 patients with asymptomatic or mild disease at the end of the observation. Older age, consolidation on chest CT, and lymphopenia on admission were more frequent in patients with severe COVID-19 (n = 28) than those with mild COVID-19 (n = 43) at the end of observation. LDH level might be marker for symptom onset in patients with COVID-19, whereas older age, consolidation on chest CT imaging, and lymphopenia are potential risk factors for disease progression. The current report findings will contribute to the improvement of clinical management in patients with COVID-19.

*Implications of all the available evidence:* Serum LDH level is a potential predictor of symptom onset of COVID-19, whereas older age, consolidation on chest CT imaging, and lymphopenia have potential utility as markers for disease progression.

## Introduction

The coronavirus disease 2019 (COVID-19) is caused by the novel coronavirus severe acute respiratory syndrome coronavirus 2 (SARS-CoV2). The first case was reported in Wuhan, China, in December 2019, ^1^ and the ongoing outbreak has been declared as a pandemic by the World Health Organization. Since specific treatment for COVID-19 is unknown, identification of patients at high risk for severe illness is critical to prepare and provide sufficient supportive therapy. Additionally, an explosive increase in case numbers has led to the collapse of healthcare systems. Therefore, triage of patients and their transportation to proper facilities are necessary. Although our knowledge of the clinical characteristics of COVID-19 and the pathogenicity of SARS-CoV2 are increasing, detailed clinical characteristics of the disease remain unknown; most of the published studies to date focus on patients with severe COVID-19. ^2-5^ In Japan, a large number of infections occurred among the passengers and crew members on board the Diamond Princess cruise ship in February 2020. By March 1, 2020, there were approximately 700 patients with laboratory-confirmed SARS-CoV2 infection. ^6,7^ Approximately 3700 passengers and crew members on the cruise ship were tested for SARS-CoV2 by reverse transcription (RT)-polymerase chain reaction (PCR), and all symptomatic and asymptomatic cases were referred to medical institutions designated for infectious diseases in accordance with the Infectious Diseases Control Law of Japan. ^6,7^ Approximately 15% of all laboratory-confirmed cases from Diamond Princess were admitted to the Self-Defense Forces Central Hospital in Japan.

In clinical settings, identifying patients who are at risk of symptom onset and clinical deterioration is essential. Therefore, understanding the clinical characteristics of patients with COVID-19 of different severity is necessary. To that end, we retrospectively analyzed the detailed clinical characteristics of hospitalized patients with COVID-19 as a result of the Diamond Princess outbreak, including those with asymptomatic, mild, and severe disease presentation, who were treated in the Self-Defense Forces Central Hospital in Japan.

## Methods

### Study design and patients

We conducted a retrospective review of the medical records of 104 patients with laboratory-confirmed COVID-19 who were admitted to the Self-Defense Forces Central Hospital in Japan between February 11 and February 25, 2020. Approximately 3700 passengers on the cruise ship were examined by quantitative RT-PCR or nested RT-PCR for SARS-CoV2 using pharyngeal swabs or sputum specimens at the public health institutes based on the recommendations protocol by the National Institute of Infectious Disease. ^6-8^ This study was reviewed and approved by the Japan Self-Defense Forces Central Hospital (approval number 01-011). Informed consent, both written and oral, was obtained from all enrolled patients.

### Data collection

Patient information was retrospectively collected from the hospital medical records and included clinical records, laboratory findings, and chest CT images. All data were collected and reviewed by two study investigators (ST and KI).

### Definitions

The geographical regions were classified according to the standard country or area codes for statistical use (M49) by the United Nations Statistics Division (https://unstats.un.org/unsd/methodology/m49/). Asymptomatic cases were defined as patients with no history of clinical signs and symptoms. Severe symptomatic cases were defined as patients showing clinical symptoms of pneumonia (dyspnea, tachypnea, peripheral capillary oxygen saturation <93%, and need for oxygen therapy). ^9^ Patients with other symptoms were classified as mild cases. The observation period was defined as the time interval between the date of admission and the date of discharge or February 26, 2020.

### Statistical analysis

Continuous variables were expressed as means with standard deviation or medians with interquartile range (IQR) and compared using Student’s *t* test and Wilcoxon rank-sum test for parametric and nonparametric data, respectively. Categorical variables were expressed as numbers (%) and compared by the χ^2^ or Fisher’s exact test. A two-sided *p* value of less than 0·05 was considered to be statistically significant. All statistical analyses were calculated using Stata13 (Stata Corp, College Station, TX, USA).

## Results

During the observation period, a total of 107 patients on board the Diamond Princess cruise ship with laboratory-confirmed COVID-19 were hospitalized in the Self-Defense Forces Central Hospital in Japan. Three patients were excluded from the study because of withdrawal of consent to join the study. Therefore, the remaining 104 patients were included in the final analysis. The patients’ clinical history, physical examination, and chest CT findings were evaluated on the day of admission, and all blood tests were completed within the first 2 days following admission. The characteristics of 104 patients are presented in Table 1. Briefly, the patient age ranged from 25 to 93 years (median, 68 years; IQR, 46·75–75 years), and 54 (51.9%) patients were male. Most of the patients were from Eastern Asia, with Japanese and Chinese ethnicities as the majority. The range of observation period was 3–15 days (median, 10 days; IQR, 7–10 days). Fifty-two (50.0%) patients had comorbidities. Based on their presentation on the day of admission, 43 patients (41.4%) who did not display any symptoms were classified as asymptomatic, whereas 41 (39.4%) and 20 (19.2%) patients were classified to have mild and severe COVID-19, respectively.

**Table 1.** Baseline characteristics of patients with COVID-19 on the Diamond Princess cruise ship.

| Total number of patients (n = 104) |  |
| --- | --- |
| **Characteristics** |  |
| Age | 68 [46·75–75] |
| Sex (male) | 54 (51·9%) |
| Observation period (days) | 10 [7–10] |
| Time from onset to admission (days) | 5 [2–7] |
| Nationality |  |
| Eastern Asia | 55 (52·9%) |
| Southeast Asia | 21 (20·2%) |
| Europe | 4 (3·8%) |
| Northern America | 14 (13·5%) |
| Oceania | 4 (3·8%) |
| Others | 6 (5·8%) |
| **Smoking** |  |
| Yes | 18 (17·3%) |
| **Comorbid condition** |  |
| Total | 52 (50·0%) |
| Cardiovascular disease | 31 (29·8%) |
| Endocrine disorder | 9 (8·7%) |
| Diabetes mellitus | 7 (6·7%) |
| Respiratory disorder | 7 (6·7%) |
| Malignancy | 4 (3·8%) |
| **Severity on admission** |  |
| Asymptomatic | 43 (41·4%) |
| Mild | 41 (39·4%) |
| Severe | 20 (19·2%) |

Data are presented as n (%), or median [interquartile range], unless otherwise specified.

The clinical findings of the patients who were symptomatic on the day of admission and at the end of observation period are shown in Table 2. At the end of the observation, there were 33 (31.7%), 43 (41.3%), and 28 (26.9%) patients with confirmed asymptomatic, mild, and severe COVID-19, respectively. The most common clinical signs and symptoms on admission among the 71 symptomatic patients at the end of the observation were fever (42.3%) and cough (40.8%). Of the 43 patients who were asymptomatic on admission, 10 (23.3%) patients developed symptomatic COVID-19 during the observation period, including three (7.0%) patients who developed severe COVID-19 and required supplemental oxygen therapy. Conversely, five of the 41 (12.2%) patients with mild disease on admission developed severe COVID-19 and required supplemental oxygen therapy. Therefore, eight of the 84 (9.5%) patients with asymptomatic or mild presentation on admission developed severe COVID-19 during the observation period. Additionally, of the 71 patients who were symptomatic at the end of the observation, 14 (13.5%) and one (1.4%) patient required oxygen therapy and mechanical ventilation, respectively, whereas 10 (14.1%) and five (7.0%) patients were treated by antibiotics and lopinavir/ritonavir, respectively. There were no reports of complications or death during the observation period.

**Table 2.** Clinical findings and disease severity of 71 patients with symptomatic COVID-19.

|  | Number (%) |  |
| --- | --- | --- |
|  | On admission | At the end of observation |
| **Signs and symptoms** |  |  |
| Fever | 30 (42.3%) | 36 (50.7%) |
| Cough | 29 (40.8%) | 43 (60.6%) |
| Malaise | 12 (16.9%) | 22 (31.0%) |
| Headache | 10 (14.1%) | 18 (25.4%) |
| Sore throat | 11 (15.5%) | 11 (15.5%) |
| Runny nose | 16 (22.5%) | 24 (33.8%) |
| Diarrhea | 8 (11.3%) | 10 (14.1%) |
| Dyspnea | 7 (9.9%) | 19 (26.8%) |
| Tachypnea | 16 (22.5%) | 24 (33.8%) |
| SpO<sub>2</sub> < 93% | 3 (4.2%) | 14 (19.7%) |
| **Treatment** |  |  |
| Acetaminophen | 4 (5.6%) | 15 (21.1%) |
| NSAIDs | - | 1 (1.4%) |
| Antibiotics | - | 10 (14.1%) |
| Lopinavir/ritonavir | - | 5 (7.0%) |
| Oxygen therapy | 3 (4.2%) | 14 (19.7%) |
| Mechanical ventilation | - | 1 (1.4%) |
| **Severity** |  |  |
| Asymptomatic | 10 (14.1%) | - |
| Mild | 41 (57.7%) | 43 (60.6%) |
| Severe | 20 (28.2%) | 28 (39.4%) |

Data are presented as n (%).
NSAID, non-steroidal anti-inflammatory drug; SpO<sub>2</sub>, peripheral capillary oxygen saturation

The characteristics of 10 patients who developed symptomatic COVID-19 during the observation period among those who were asymptomatic on admission are presented in Table 3. Among these 10 patients who developed symptomatic COVID-19, the most frequent symptom was cough (n = 7, 70.0%). Conversely, 17 of the 33 (51.5%) patients never developed symptoms until the end of the observation period had abnormal lung findings with various patterns on CT images (Table 3 and Figure 1). By univariate analysis, there were no significant differences in the distribution of age, sex, and comorbidities or the prevalence of abnormal lung findings and CT patterns between the symptomatic and asymptomatic patients. The prevalence of elevated LDH levels above 230 IU/L was significantly higher in patients who were initially asymptomatic but developed symptomatic COVID-19 during the observation period (50.0% vs 12.1%; odds ratio [OR], 7.25; 95% confidence interval [CI], 1.43–36.70; *p* = 0.02).

**Table 3.** Clinical characteristics of 43 asymptomatic patients on admission.

|  | Total<br>n = 43 | At the end of observation |  | *p* value | OR<br>(95% CI) |
| --- | --- | --- | --- | --- | --- |
|  |  | Asymptomatic<br>n = 33 | Symptomatic<br>n = 10 |  |  |
| Characteristics |  |  |  |  |  |
| Age | 69<br>[60.5–75] | 70<br>[57–75] | 67.5<br>[66.25–73.5] | 0.96 |  |
| Sex (male) | 19 (44.2%) | 15 (45.5%) | 4 (40.0%) | 1.00 |  |
| Smoking |  |  |  |  |  |
| Yes | 7 (16.3%) | 5 (15.2%) | 2 (20.0%) | 0.65 |  |
| Comorbid condition |  |  |  |  |  |
| Total | 21 (48.8%) | 15 (45.5%) | 6 (60.0%) | 0.48 |  |
| Cardiovascular disease | 12 (27.9%) | 9 (27.3%) | 3 (30.0%) | 1.00 |  |
| Endocrine disorder | 3 (7.0%) | 2 (6.1%) | 1 (10.0%) | 0.56 |  |
| Diabetes mellitus | 5 (11.6%) | 5 (15.2%) | - | 0.32 |  |
| Respiratory disorder | 4 (9.3%) | 2 (6.1%) | 2 (20.0%) | 0.22 |  |
| Malignancy | 2 (4.7%) | 1 (3.0%) | 1 (10.0%) | 0.42 |  |
| Signs and symptoms |  |  |  |  |  |
| Fever | 3 (7.0%) | - | 3 (30.0%) | - |  |
| Cough | 7 (16.3%) | - | 7 (70.0%) | - |  |
| Malaise | 2 (4.7%) | - | 2 (20.0%) | - |  |
| Headache | 2 (4.7%) | - | 2 (20.0%) | - |  |
| Sore throat | - | - | - | - |  |
| Runny nose | - | - | - | - |  |
| Diarrhea | 1 (2.3%) | - | 1 (10.0%) | - |  |
| Dyspnea | 3 (7.0%) | - | 3 (30.0%) | - |  |
| Tachypnea | 3 (7.0%) | - | 3 (30.0%) | - |  |
| SpO<sub>2</sub> < 93% | 3 (7.0%) | - | 3 (30.0%) | - |  |
| Radiographically findings on admission |  |  |  |  |  |
| Abnormal lung findings | 25 (58.1%) | 17 (51.5%) | 8 (80.0%) | 0.15 |  |
| Lateral | 13 (30.2%) | 10 (30.3%) | 3 (30.0%) | - |  |
| Bilateral | 12 (27.9%) | 7 (21.2%) | 5 (50.0%) | 0.11 |  |
| GGO | 15 (34.9%) | 11 (33.3%) | 4 (40.0%) | 0.72 |  |
| Multifocal GGO | 12 (27.9%) | 9 (27.3%) | 3 (30.0%) | 1.00 |  |
| Consolidation | 9 (20.9%) | 6 (18.2%) | 3 (30.0%) | 0.41 |  |
| Crazy-paving appearance | 5 (11.6%) | 2 (6.1%) | 3 (30.0%) | 0.07 |  |
| Nodular lesion | 3 (7.0%) | 2 (6.1%) | 1 (10.0%) | 0.56 |  |
| **Laboratory findings on admission** |  |  |  |  |  |
| AST > 38 (IU/L) | 4 (9.3%) | 3 (9.1%) | 1 (10.0%) | 1.00 |  |
| ALT > 45 (IU/L) | 5 (11.6%) | 4 (12.1%) | 1 (10.0%) | 1.00 |  |
| LDH > 230 (IU/L) | 9 (20.9%) | 4 (12.1%) | 5 (50.0%) | 0.02* | 7.25<br>(1.43–36.70) |
| CRP > 1.0 (mg/dL) | 18 (41.9%) | 11 (33.3%) | 7 (70.0%) | 0.07 |  |
| White blood cell count<br><4000 (/μL) | 6 (14.0%) | 4 (12.1%) | 2 (20.0%) | 0.61 |  |
| Platelet count<br><15 (×10<sup>4</sup>/μL) | 2 (4.7%) | 1 (3.0%) | 1 (10.0%) | 0.41 |  |
| Lymphocyte count<br><1200 (/μL) | 13 (30.2%) | 9 (27.3%) | 4 (40.0%) | 0.46 |  |

Data are presented as n (%), or median [interquartile range], unless otherwise specified.
Asterisk indicates statistical significance.
OR, odds ratio; 95% CI, 95% confidence interval; GGO, ground glass opacity; AST, aspartate aminotransferase; ALT, alanine aminotransferase; LDH, lactate dehydrogenase; CRP, C reactive protein

**Figure 1.**
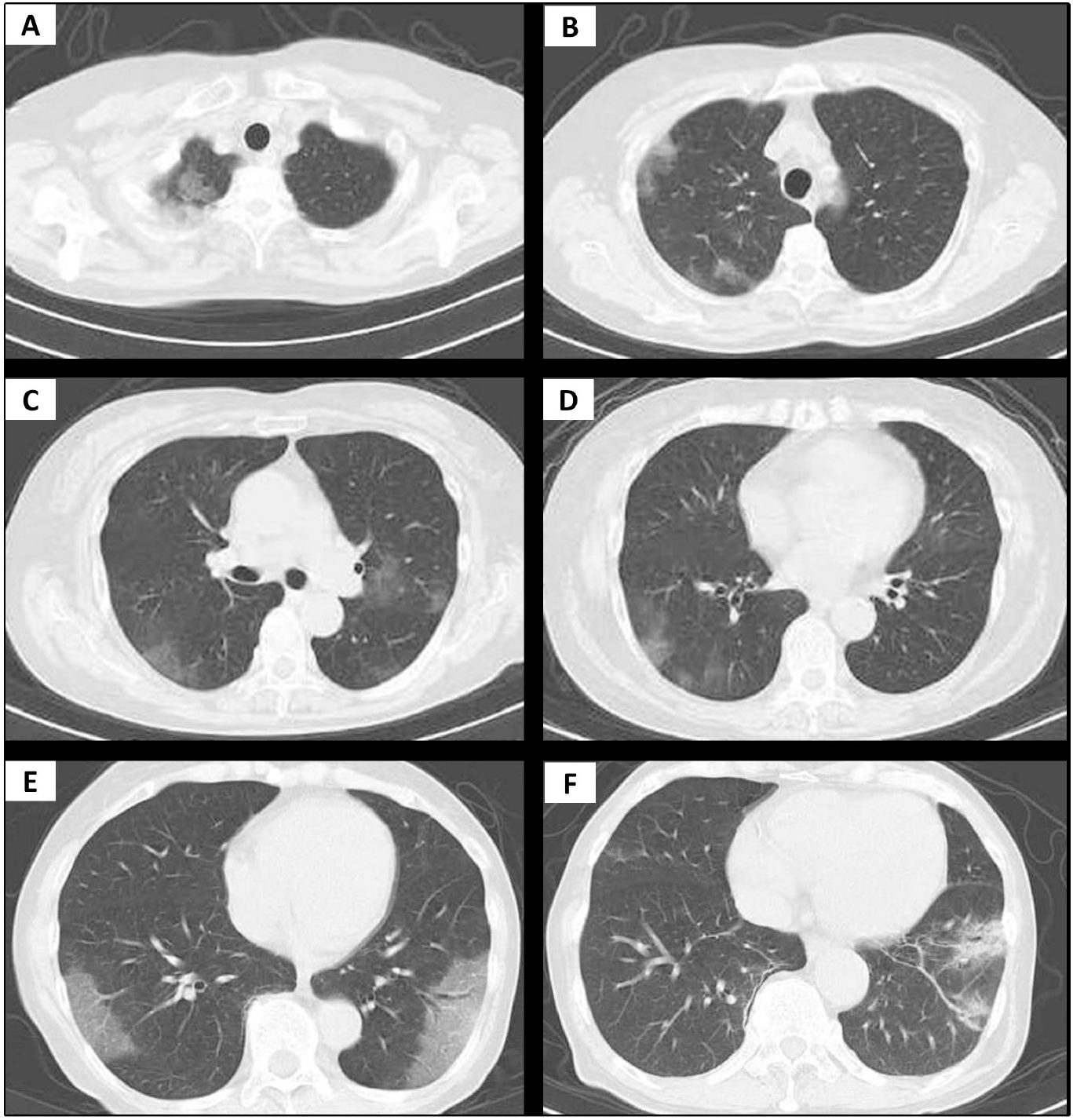
Computed tomography (CT) patterns of abnormal lung findings among asymptomatic patients with the 2019 novel coronavirus disease (COVID-19). (A–D) A 73 year-old female patient. Patchy non-segmental GGOs are observed adjacent to the parietal pleura in the right upper lobe (A and B) and in both lower lobes (C and D). (E) A 70 year-old female patient. GGO with interlobular septal thickening (crazy-paving appearance) adjacent to the parietal pleura in both lower lobes. (F) A 76 year-old male. CT image shows GGO with consolidation, bronchial wall thickening, and bronchiectasis in left lower lobe.

In the group of eight patients who developed severe COVID-19 during the observation period, the age range was 42–87 years (median, 70 years; IQR, 70–77 years), and five (62.5%) patients were male. The period from symptom onset to the reclassification of disease presentation as severe COVID-19 ranged from 1 to 5 days (median, 4 days; IQR, 2.5–5 days). Additionally, all eight patients (100.0%) developed radiographical abnormalities on CT prior to the emergence of symptoms of typical pneumonia including tachypnea, dyspnea, and hypoxemia (Figure 2). Furthermore, among these patients who developed severe COVID-19 during the observation period, 4 (50.0%) and 3 (37.5%) patients lacked the symptoms of cough and fever, respectively. We also compared these eight patients who developed severe COVID-19 with 76 patients who remained asymptomatic or had mild COVID-19 at the end of the observation period. By univariate analysis, no significant differences were observed in the distribution of age, sex, and comorbidities between the two groups. The prevalence rates of consolidation (75.0% vs 19.7%; OR, 12.20; 95%CI, 2.23–66.59; *p* = 0.04) and lymphopenia (62.5% vs 25.0%; OR, 5.00; 95%CI, 1.10–22.92; *p* = 0.04) on the day of admission were significantly higher in patients who developed severe COVID-19 during the observation period than in those who were asymptomatic or had mild disease at the end of the observation period.

**Figure 2.**
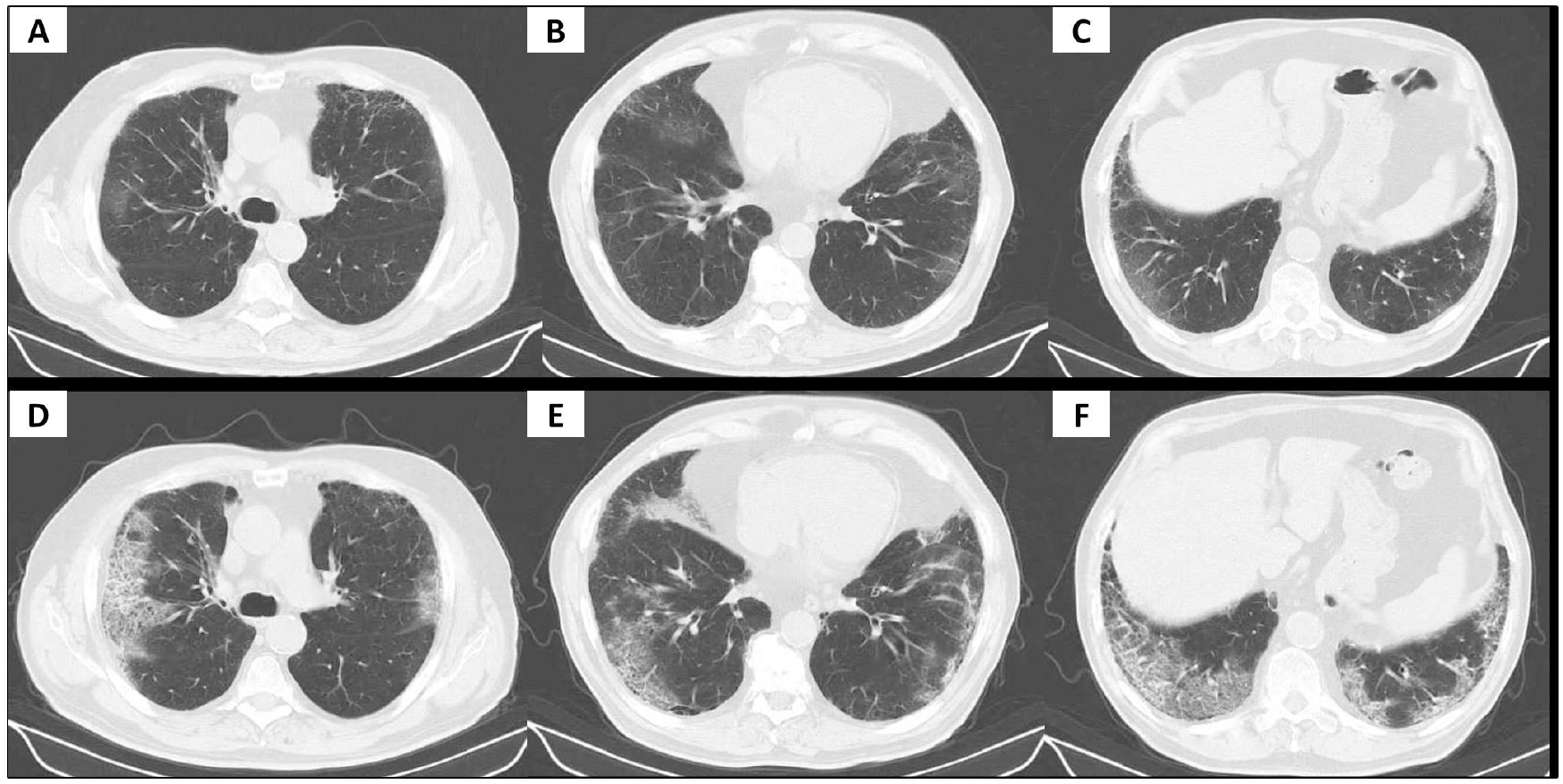
Progression of CT findings in a 75 year-old male patient with COVID-19. The patient was asymptomatic on the day of admission. On the fourth day of admission, he developed tachypnea and hypoxemia and was administered oxygen therapy. (A–C) Chest CT images on the day of admission show multifocal GGO adjacent to the parietal pleura in multiple lobes with emphysematous changes. (D–F) Follow-up chest CT images on the tenth day of admission show an increase in the extent of GGO with crazy-paving appearance.

Finally, we performed analyses to identify differences in the clinical characteristics between patients with mild and severe COVID-19 at the end of the observation period (Table 4). By univariate analysis, age was significantly higher in the severe COVID-19 group than in the mild COVID-19 group (median, 72.5 years; IQR, 55.25–76.5 years vs median, 60 years; IQR, 40.0 –71.0 years; *p* = 0.03). There were no significant differences in the distribution of sex, comorbidities, clinical signs, and symptoms between the two groups. However, there was a significant difference in the prevalence of consolidation as a CT pattern between the severe and mild cases (46.4% vs 20.9%; OR, 3.27; 95%CI, 1.15–9.30; *p* = 0.03). Among the blood tests, the rate of lymphopenia was higher in patients with severe COVID-19 than in those with mild COVID-19 at the end of the observation period (57.1% vs 23.3%; OR, 4.40; 95% CI; 1.57–12.32; *p* = 0.005).

**Table 4.**
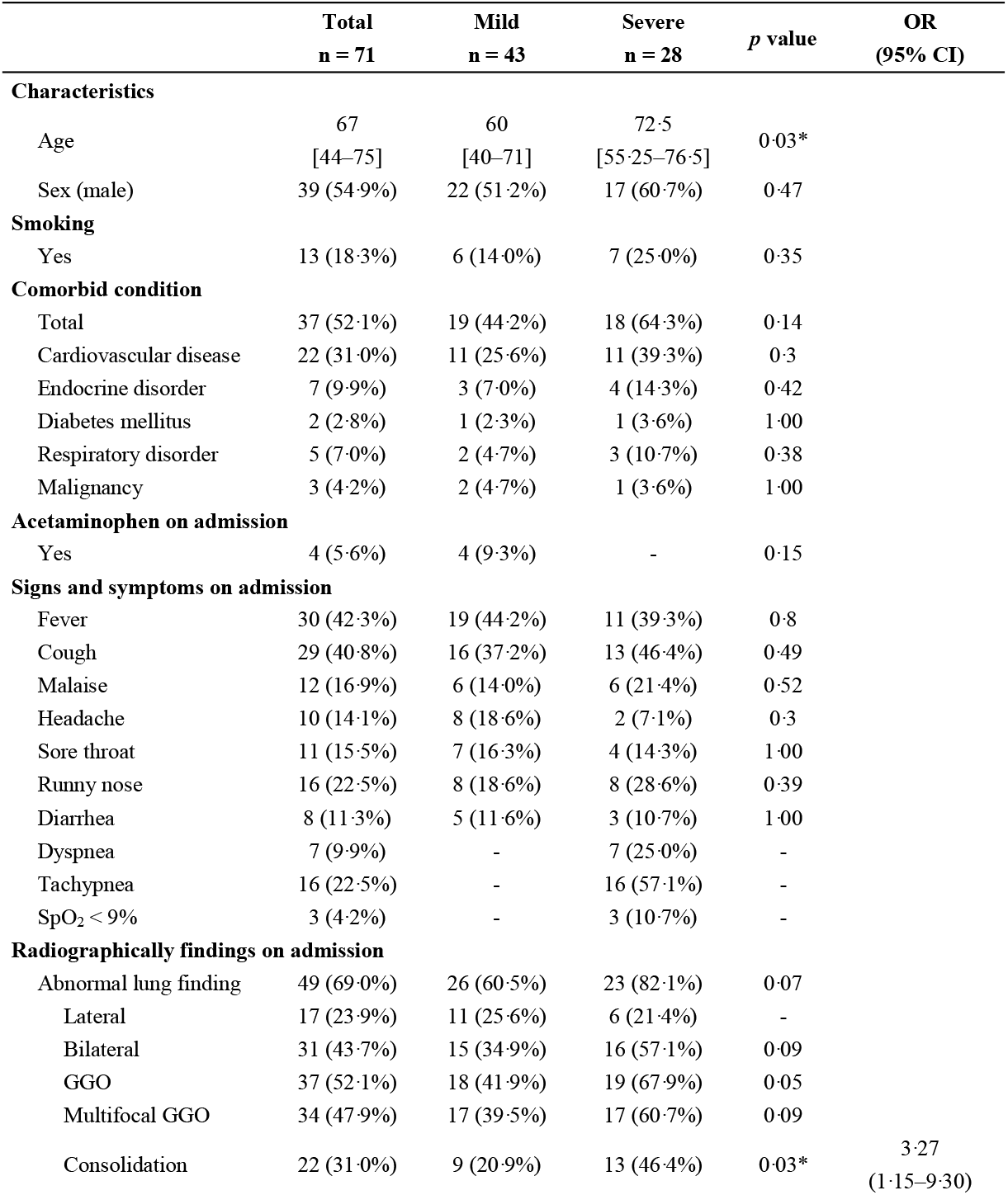
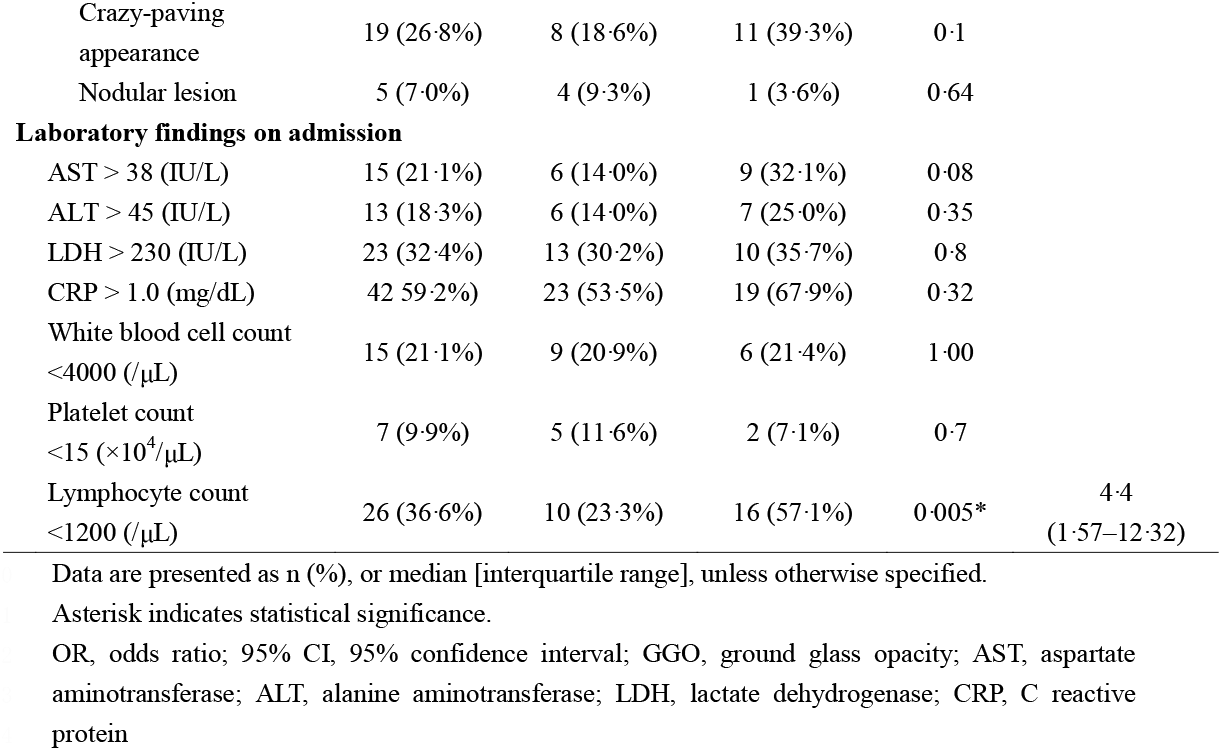
The clinical characteristics of patients with mild and severe COVID-19 at the end of observation.

|  | **Total**<br>**n = 71** | **Mild**<br>**n = 43** | **Severe**<br>**n = 28** | ***p* value** | **OR**<br>**(95% CI)** |
| --- | --- | --- | --- | --- | --- |
| **Characteristics** |  |  |  |  |  |
| Age | 67<br>[44–75] | 60<br>[40–71] | 72.5<br>[55.25–76.5] | 0.03* |  |
| Sex (male) | 39 (54.9%) | 22 (51.2%) | 17 (60.7%) | 0.47 |  |
| **Smoking** |  |  |  |  |  |
| Yes | 13 (18.3%) | 6 (14.0%) | 7 (25.0%) | 0.35 |  |
| **Comorbid condition** |  |  |  |  |  |
| Total | 37 (52.1%) | 19 (44.2%) | 18 (64.3%) | 0.14 |  |
| Cardiovascular disease | 22 (31.0%) | 11 (25.6%) | 11 (39.3%) | 0.3 |  |
| Endocrine disorder | 7 (9.9%) | 3 (7.0%) | 4 (14.3%) | 0.42 |  |
| Diabetes mellitus | 2 (2.8%) | 1 (2.3%) | 1 (3.6%) | 1.00 |  |
| Respiratory disorder | 5 (7.0%) | 2 (4.7%) | 3 (10.7%) | 0.38 |  |
| Malignancy | 3 (4.2%) | 2 (4.7%) | 1 (3.6%) | 1.00 |  |
| **Acetaminophen on admission** |  |  |  |  |  |
| Yes | 4 (5.6%) | 4 (9.3%) | - | 0.15 |  |
| **Signs and symptoms on admission** |  |  |  |  |  |
| Fever | 30 (42.3%) | 19 (44.2%) | 11 (39.3%) | 0.8 |  |
| Cough | 29 (40.8%) | 16 (37.2%) | 13 (46.4%) | 0.49 |  |
| Malaise | 12 (16.9%) | 6 (14.0%) | 6 (21.4%) | 0.52 |  |
| Headache | 10 (14.1%) | 8 (18.6%) | 2 (7.1%) | 0.3 |  |
| Sore throat | 11 (15.5%) | 7 (16.3%) | 4 (14.3%) | 1.00 |  |
| Runny nose | 16 (22.5%) | 8 (18.6%) | 8 (28.6%) | 0.39 |  |
| Diarrhea | 8 (11.3%) | 5 (11.6%) | 3 (10.7%) | 1.00 |  |
| Dyspnea | 7 (9.9%) | - | 7 (25.0%) | - |  |
| Tachypnea | 16 (22.5%) | - | 16 (57.1%) | - |  |
| SpO<sub>2</sub> < 9% | 3 (4.2%) | - | 3 (10.7%) | - |  |
| **Radiographically findings on admission** |  |  |  |  |  |
| Abnormal lung finding | 49 (69.0%) | 26 (60.5%) | 23 (82.1%) | 0.07 |  |
| Lateral | 17 (23.9%) | 11 (25.6%) | 6 (21.4%) | - |  |
| Bilateral | 31 (43.7%) | 15 (34.9%) | 16 (57.1%) | 0.09 |  |
| GGO | 37 (52.1%) | 18 (41.9%) | 19 (67.9%) | 0.05 |  |
| Multifocal GGO | 34 (47.9%) | 17 (39.5%) | 17 (60.7%) | 0.09 |  |
| Consolidation | 22 (31.0%) | 9 (20.9%) | 13 (46.4%) | 0.03* | 3.27<br>(1.15–9.30) |

|  |  |  |  |  |
| --- | --- | --- | --- | --- |
| Crazy-paving appearance | 19 (26.8%) | 8 (18.6%) | 11 (39.3%) | 0.1 |
| Nodular lesion | 5 (7.0%) | 4 (9.3%) | 1 (3.6%) | 0.64 |
| **Laboratory findings on admission** |  |  |  |  |  |
| AST > 38 (IU/L) | 15 (21.1%) | 6 (14.0%) | 9 (32.1%) | 0.08 |  |
| ALT > 45 (IU/L) | 13 (18.3%) | 6 (14.0%) | 7 (25.0%) | 0.35 |  |
| LDH > 230 (IU/L) | 23 (32.4%) | 13 (30.2%) | 10 (35.7%) | 0.8 |  |
| CRP > 1.0 (mg/dL) | 42 (59.2%) | 23 (53.5%) | 19 (67.9%) | 0.32 |  |
| White blood cell count <4000 (/μL) | 15 (21.1%) | 9 (20.9%) | 6 (21.4%) | 1.00 |  |
| Platelet count <15 (×10<sup>4</sup>/μL) | 7 (9.9%) | 5 (11.6%) | 2 (7.1%) | 0.7 |  |
| Lymphocyte count <1200 (/μL) | 26 (36.6%) | 10 (23.3%) | 16 (57.1%) | 0.005* | 4.4<br>(1.57–12.32) |

Data are presented as n (%), or median [interquartile range], unless otherwise specified.
Asterisk indicates statistical significance.
OR, odds ratio; 95% CI, 95% confidence interval; GGO, ground glass opacity; AST, aspartate aminotransferase; ALT, alanine aminotransferase; LDH, lactate dehydrogenase; CRP, C reactive protein

## Discussion

The present retrospective, single-center study provides a summary of the clinical features of patients who developed COVID-19 of varying severity as a result of mass infection on board a cruise ship. The LDH levels were significantly higher in 10 patients who were initially asymptomatic on admission to the hospital and developed symptomatic COVID-19 during the observation period than in 33 patients who remained asymptomatic throughout the observation period. Older age, consolidation on chest CT images, and lymphopenia at the time of admission were significantly more frequent among 28 patients with severe COVID-19 than among 43 patients with mild COVID-19 at the end of the observation period. Overall, these findings suggest that serum LDH level might be a marker of symptom onset and that older age, consolidation on chest CT images, and lymphopenia are potential risk factors for disease progression in patients with COVID-19.

The predictors of symptom onset and risk factors for disease progression in COVID-19 remain partially unclear because patients who are asymptomatic or are otherwise exhibiting mild symptoms do not necessarily visit a hospital. During the COVID-19 outbreak on the Diamond Princess cruise ship, approximately 3700 individuals on board were tested for SARS-CoV2 by RT-PCR, and all laboratory-confirmed patients were referred to the hospitals. This unique situation allowed us to observe the clinical course of asymptomatic patients as well as those with mild and severe symptoms, which has not been reported to date. ^2-5^

Based on the previously published findings of COVID-19 in China, old age, male sex, and presence of comorbidities are considered as potential risk factors for disease progression and poor prognosis. ^1,10^ Zhou *et al*. indicated that older age, high sequential organ failure assessment score, and elevated d-dimer level were potential risk factors for death in patients with COVID-19. ^11^ Qin *et al*. reported that lymphopenia and higher neutrophil–lymphocyte ratio were frequently observed in patients with severe COVID-19 compared with those with mild disease. The present study findings support those of previous reports suggesting older age and lymphopenia as potential risk factors for disease progression in COVID-19. Lymphopenia, a common feature of symptomatic COVID-19, ^12,13^ can be a direct consequence of SARS-CoV2 or might be mediated by cytokines. Qin *et al*. also suggested that SARS-CoV2 infection might affect T lymphocytes and dysregulate the immune system, consequently leading to secondary bacterial infections. ^14^ In the present study, we observed lymphopenia in 27.3% of the patients who were asymptomatic at the end of observation period, which suggests the development of immune system dysregulation as a direct consequence of SARS-CoV2 infection in asymptomatic cases. However, the role lymphopenia in asymptomatic patients remains unclear. Further cohort studies are warranted to determine the effect of lymphopenia caused by SARS-CoV2 infection in the presence of secondary bacterial infection.

In the present study, the prevalence of consolidation detected by chest CT was significantly higher in severe cases than in mild cases. Consolidation was also frequently detected in patients who developed severe COVID-19 during the observation period. Pan *et al*. and Shi *et al*. reported that abnormal lung findings varied from ground glass opacities (GGOs) to consolidation during the course of COVID-19 and that the appearance of consolidation was more frequent within 1–3 weeks after the symptom onset. ^15,16^ Previous studies found that 55% of patients with COVID-19 developed dyspnea in a median of 8 days (IQR, 5.0–13.0 days) after symptom onset; ^1^ thus, the development of consolidation on CT is associated with disease deterioration. Generally, consolidation is easily detectable by chest X-ray, which therefore might be useful for the evaluation of risk for disease progression in patients with COVID-19 in settings with limited resources.

Additionally, CT imaging revealed a high prevalence of lung abnormalities in patients with COVID-19. The various patterns of abnormal lung findings were observed in asymptomatic cases (51.5%) as well as in symptomatic cases (69.0%) at the end of the observation period. In their report focusing on CT imaging findings of COVID-19, Shi *et al*. also noted the presence of abnormal lung findings in asymptomatic patients. ^16^ Bai *et al*. investigated the common radiological characteristics of COVID-19 pneumonia and reported that chest CT had a diagnostic sensitivity of 73%–93% and a specificity of 93%–100% in distinguishing COVID-19 from viral pneumonia. ^17^ Therefore, chest CT imaging might also be useful diagnostic tool for the asymptomatic and symptomatic patients with COVID-19. In the present study, all eight patients who developed severe COVID-19 during the observation period presented with CT abnormalities, in advance of the appearance of typical pneumonia symptoms such as dyspnea, tachypnea, and hypoxemia. Of these eight patients, four and three patients lacked the symptoms of cough and fever, respectively. These findings highlight that COVID-19 pneumonia silently progresses without any remarkable symptoms. Conversely, the majority of asymptomatic patients with radiological abnormalities on CT imaging did not develop severe pneumonia. The differences observed in the clinical outcomes might be associated with patient background characteristics as well as the genetic and pathogenic diversity of SARS-CoV2. Understanding CT abnormalities relevance to the duration of virus shedding and the incidence of secondary bacterial pneumonia is important to improve the clinical management of COVID-19. Further investigation is necessary to clarify the clinical impact of CT findings in patients with asymptomatic or mild COVID-19.

Interestingly, there was a significant difference in the rate of patients with elevated LDH levels between those who developed symptomatic COVID-19 during the observation period and those who remained asymptomatic throughout the observation period, although there was no significant difference in the prevalence of lymphopenia between the two groups. Elevated serum LDH levels are associated with lung tissue damage, ^18^ and serum LDH is considered as a marker for disease activity and progression in pneumonia due to various etiologies. ^19,20^ Given that abnormal lung findings are frequently observed in patients with asymptomatic COVID-19, these findings implicate that an elevated LDH level might also reflect lung tissue damage by COVID-19 and might be considered as a predictor for symptom onset in COVID-19. Patient assessment with determination of serum LDH levels and chest CT imaging might facilitate the successful dedication of medical resources to patients who are at higher risk for disease progression in the setting of mass infections as well as in normal clinical settings.

The major limitations of the present study were selection bias and low number of patients. We enrolled an ideal, homogeneous cluster of relatively healthy individuals who could remain on board a cruise ship for a relatively long term. Therefore, the patient population was not an exact representation of normal clinical settings. Triage on the Diamond Princess cruise ship before emergency transportation has also introduced a selection bias with an atypically large number of patients. The Self-Defense Forces Central Hospital in Tokyo is close to the Yokohama port where the cruise ship was docked, and the symptomatic cases were referred to the study hospital. However, the critical cases were preferentially referred to neighboring hospitals in Yokohama. Likewise, several clinical signs and symptoms, which have occurred and disappeared before the patients were referred to our hospital, may be included or excluded from this study. In addition, laboratory tests and CT images were obtained once in majority of the patients, and the present study was not able to determine changes in lymphocyte count and radiological findings over the course of observation. Therefore, multicenter, multi-national cohort studies should be conducted to determine predictive markers for symptom onset and risk factors for diseases progression.

## Conclusion

The present study highlights the clinical features of patients who developed COVID-19 as a result of mass infection on board the Diamond Princess cruise ship. Our analyses reveal serum LDH level as a potential predictor of symptom onset in COVID-19. Age, consolidation on chest CT images, and lymphopenia might be risk factors for disease progression. These findings contribute to a better understanding of the clinical course and access points to improve disease management in patients with COVID-19. Additional cohort studies are warranted to determine predictors for symptom onset and risk factors for disease progression.

## Data Availability

The datasets generated during and/or analysed during the current study are available from the corresponding author on reasonable request.

## Authors’ contributions

TI and KT, study conception and design; ST and KI, collecting data and performing data analysis; ST, KI, MI, and KM manuscript drafting and editing; TaK, HO, SK, TsK, MK and SM, manuscript revision; MS, SS and UY, study supervision. All authors have read and approved the final manuscript.

## Acknowledgments

We thank everyone involved in the COVID-19 management and treatment team from the Self-Defense Forces Central Hospital in Japan and members who were assembled from other institutes of Japan Self-Defense Forces. Particularly, we thank Dr. Koji Kameda, Dr. Takayuki Yamamoto, Dr. Daishi Higashiyama, Dr. Yoshitaka Imoto, Dr. Masataka Miyama, Dr. Tsukasa Mizuno, Dr. Kento Kato, Dr. Mamoru Honda, Dr. Shoji Takeda, and Dr. Masumi Ogawa, Dr. Shingo Tanaka, Dr. Hisashi Sasaki, Dr. Mitsuki Yamaga, Dr. Shinichiro Ota and Dr. Hiroaki Taniguchi for supporting the data collection, and Dr. Akira Fujikawa and Dr. Shohei Inui for the interpretation of chest CT images. We also thank Mr. Shingo Tamaki (School of Tropical Medicine and Global Health, Nagasaki University, Japan) for the supporting statistical analysis, Colonel. Kazuhiro Nakaya (Medical Department, Ground Staff Office, Ministry of Defense) for the scientific advice, and Enago (www.enago.jp) for the English language review.

